# Genetic variants in UNC93B1 predispose to childhood-onset systemic lupus erythematosus

**DOI:** 10.1101/2023.10.15.23296507

**Authors:** Mahmoud Al-Azab, Elina Idiiatullina, Meng Lin, Katja Hrovat-Schaale, Huifang Xian, Jianheng Zhu, Mandy Yang, Bingtai Lu, Ziyang Liu, Zhiyao Zhao, Yiyi Liu, Jingjie Chang, Xiaotian Li, Caiqin Guo, Ping Zeng, Jun Cui, Xia Gao, Yan Zhang, Yuxia Zhang, Seth L. Masters

**Author notes:** These authors contributed equally (Yuxia Zhang, Seth L. Masters). Corresponding authors: Prof Seth Masters, Prof Yuxia Zhang.

## Abstract

Rare genetic variants in TLR7 are known to cause lupus in humans and mice. UNC93B1 is a transmembrane protein that regulates TLR7 localisation into endosomes, however it has only been genetically linked to lupus in mice and dogs. We now identify a novel variant in UNC93B1 (T314A) located proximally to the TLR7 transmembrane domain, in a patient with childhood-onset systemic lupus erythematosus (SLE). Further examination in a cohort of East Asian patients revealed seven who encode UNC93B1 (V117L), which is a rare but highly significant risk factor for this disease, when compared to the corresponding general population. The variant is associated with increased expression of type I IFNs and NF-kB cytokines in patients plasma and PBMC. This was confirmed using cell line models, with exaggerated responses to stimulation of TLR7/8, but not TLR3 or TLR9, and the process can be inhibited by targeting the TLR signaling molecules IRAK1/4. For UNC93B1 (V117L) we then created the orthologous mutation in mice (V138L), which results in a spontaneous lupus-like disease in heterozygotes, that is more severe in homozygotes and again hyperresponsive to TLR7 stimulation. Together, this work formally identifies genetic variants in UNC93B1 that can predispose to childhood-onset SLE.

**One sentence summary:** Rare genetic variants in UNC93B1 predispose to childhood-onset lupus via TLR7-IRAK1/4, validated with a corresponding mouse model.

## Introduction

Systemic lupus erythematosus (SLE) is a chronic autoimmune disease that typically develops in adults, but can affect around 1/100,000 children ^1^. There is a significant genetic contribution to the condition, ranging from common variants with small effects, through to fully penetrant disease-causing alleles ^2^. One example of this is TLR7, which marks a genetic interval that is a risk factor to develop SLE ^3^, but can also drive a monogenic form of the disease due to gain-of-function mutations ^4^. TLR7 typically functions as a sensor of viral ssRNA in endosomes, to which it is trafficked by the transmembrane protein UNC93B1 ^5^. TLR7 then signals through Myd88 to IRAK1/4 leading to NF-kB and type I IFN expression programs that are typically associated with SLE ^6^, but critically required to fight viral infection. Consequently, people with loss-of-function mutations in TLR7 or UNC93B1 are immunodeficient ^7,8^. On the other hand, again similar to TLR7, UNC93B1 expression is increased in SLE patients with active disease ^9^, and mutations in murine UNC93B1 can cause a lupus-like disease ^10,11^. There is also a sporadic lupus-like disease in dogs that is due to a mutation in UNC93B1 ^12^. Despite all of this compelling data linking UNC93B1 to disease, there was still no genetic variant in the human population that formally validated its role in SLE pathogenesis.

## Identification of rare UNC93B1 variants in patients with childhood-onset SLE

Given the recent observation that rare genetic variants in TLR7 can cause childhood-onset SLE ^4^, we searched in this patient population for variants which may influence the interaction between TLR7 and the transmembrane (TM) protein which regulates its cellular localization, UNC93B1 ^13^. We first identified the novel variant UNC93B1 (c.A940G p.T314A), which is not present in the general population, and is highly conserved (**Fig. 1a**). Based on a published structure of UNC93B1 in complex with TLR7 ^13^, the affected amino acid is at the start of helix 2 (H2) between TM helices TM6 and TM7 of UNC93B1 (**Fig. 1b**). H2 is positioned on top of TM helix of TLR7 responsible for the UNC93B1-TLR7 interaction. Additionally, T314 is in close proximity to the C-terminus of the protein, a disordered region that has been shown to have two phosphorylation sites (S547 and S550) that regulate TLR7 activation ^11^. Looking more broadly in a cohort of 272 patients with pediatric lupus recruited at the Guangzhou Women’s and Children’s Medical Centre, we found that 7 encode UNC93B1 (c.G349T p.V117L) which lies at the interface between UNC93B1 protomers in the reported structure (**Fig. 1b**). Although these protomers are close to one another, there are no noticeable interactions that would prevent dimerisation of TLR7, similar to what occurs for TLR3 ^13^. Instead, V117L may act in a similar way to K333R, which is not proximal, but is also located at an UNC93B1 protomer interface and results in increased TLR7 activation. However as K333 is ubiquitinated this could represent a different mechanism of activation ^11^. The highly conserved UNC93B1 V117L variant (**Fig. 1a**) is present in the general East Asian population, and is most prevalent in South Coast Han (**Fig. 1c**) ^14^. We calculate that it confers a 17.9 fold increased risk of developing childhood-onset SLE (**Supp Table 1**). For the seven SLE patients identified to carry this allele, there were no immediate reports of affected family members, although follow-up studies would be required to confirm this.

**Fig. 1.**
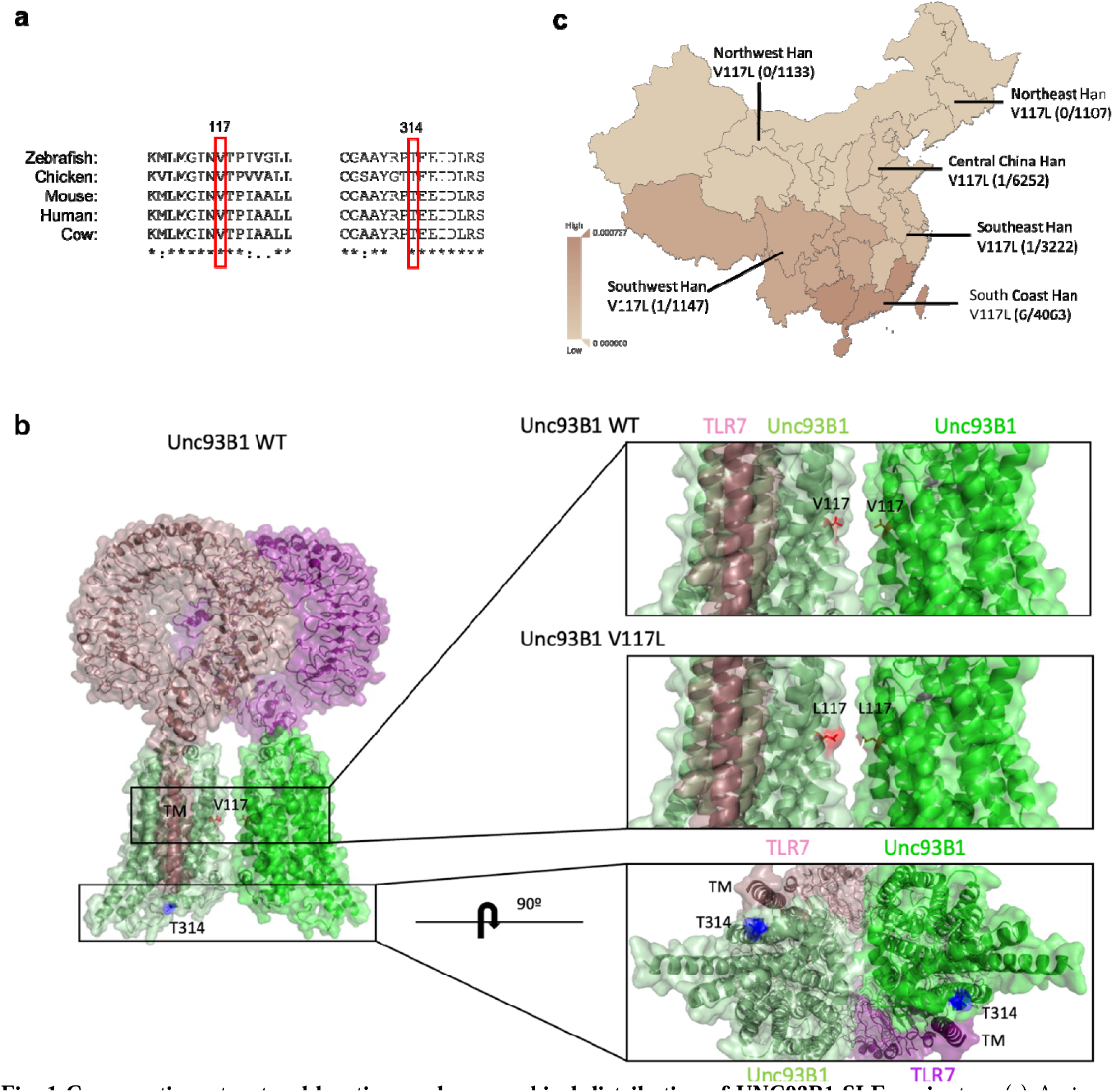
Conservation, structural location, and geographical distribution of UNC93B1 SLE variants. (a) Amino acid conservation across 5 species, as indicated, for residues surrounding the variants of interest in UNC93B1. (b) UNC93B1 (PDB:7cyn) shown in light green and green (surface and cartoon representation) with T314 shown in blue sticks on H2. TLR7 protomers are shown in purple and pink (surface and cartoon representation). V117 is shown in red sticks between the interface of two UNC93B1 protomers, L117 is shown in magenta. (c) UNC93B1 V117L variant local geographical distribution.

Clinical parameters for patients encoding UNC93B1 V117L (P1, P2, P3) and T314A (P4) include elevated titers of autoantibodies to double stranded deoxyribonucleic acid (anti-dsDNA) and anti-nuclear antibodies (ANA), with increased protein in 24h collected urine and depleted serum complement 3/4 (**Fig. 2a and Supp Fig. 1a-c**). Comprehensive light microscopy, immunofluorescence and electron microscopy studies for P4 are consistent with diffuse proliferative lupus nephritis with membranous lupus nephritis, IV-G (A) + V (**Fig. 2b-f**). Complete blood count for P2, P3, and P4 were generally normal, with periods of neutrophilia, corresponding to periods of lymphopenia (**Supp Fig. 1a**). The overall disease score was clinically evaluated using SLEDAI and SLEDAI-2k scoring systems where results ranged from 2 to 20 (**Fig. 2a and Supp Fig. 1a**). Other clinical parameters and treatments are reported in **Supp Table 2**. We isolated UNC93B1 (V117L) patients, P2 and P3, peripheral blood mononuclear cells (PBMCs) and found spontaneous expression of IL-8, MX1, and ISG-15 genes in P2 (**Fig. 2g and Supp Fig. 2a**), and IL-8, MX1, IFIT1, IFIT3, and ISG-15 in P3 (**Fig. 2h and Supp Fig. 2b**). Indeed, we observed induced secretion of typical TLR7 induced cytokines, IL-8, IP-10, IFNγ, IFN-λ1/2/3, IL-12p70, IL-10, IFNα, IFNβ and IL-6, but not TNFα in P2 (**Fig. 2i and Supp Fig. 2c, d**), and IL-8, IFNα, IFNγ, IFN-λ1/2/3, IL-10, and IL-12p70 in P3 (**Fig. 2j and Supp Fig. 2e**). Furthermore, we detected elevated levels of IP-10, IL-8, and TNFα in plasma of P2 and P3 (**Fig. 2k),** and IFN-α2, IFNγ, IFN-λ1/2/3, IL-6, IL-12p70, IL-1β, GM-CSF, and IL-10 in UNC93B1 (V117L) P2 plasma (**Supp Fig. 2f**). These data identify rare UNC93B1 variants in patients with childhood-onset SLE, associated with typical clinical parameters and elevated levels of lupus-associated cytokines, similar to patients with mutations in TLR7 ^4^.

**Fig. 2.**
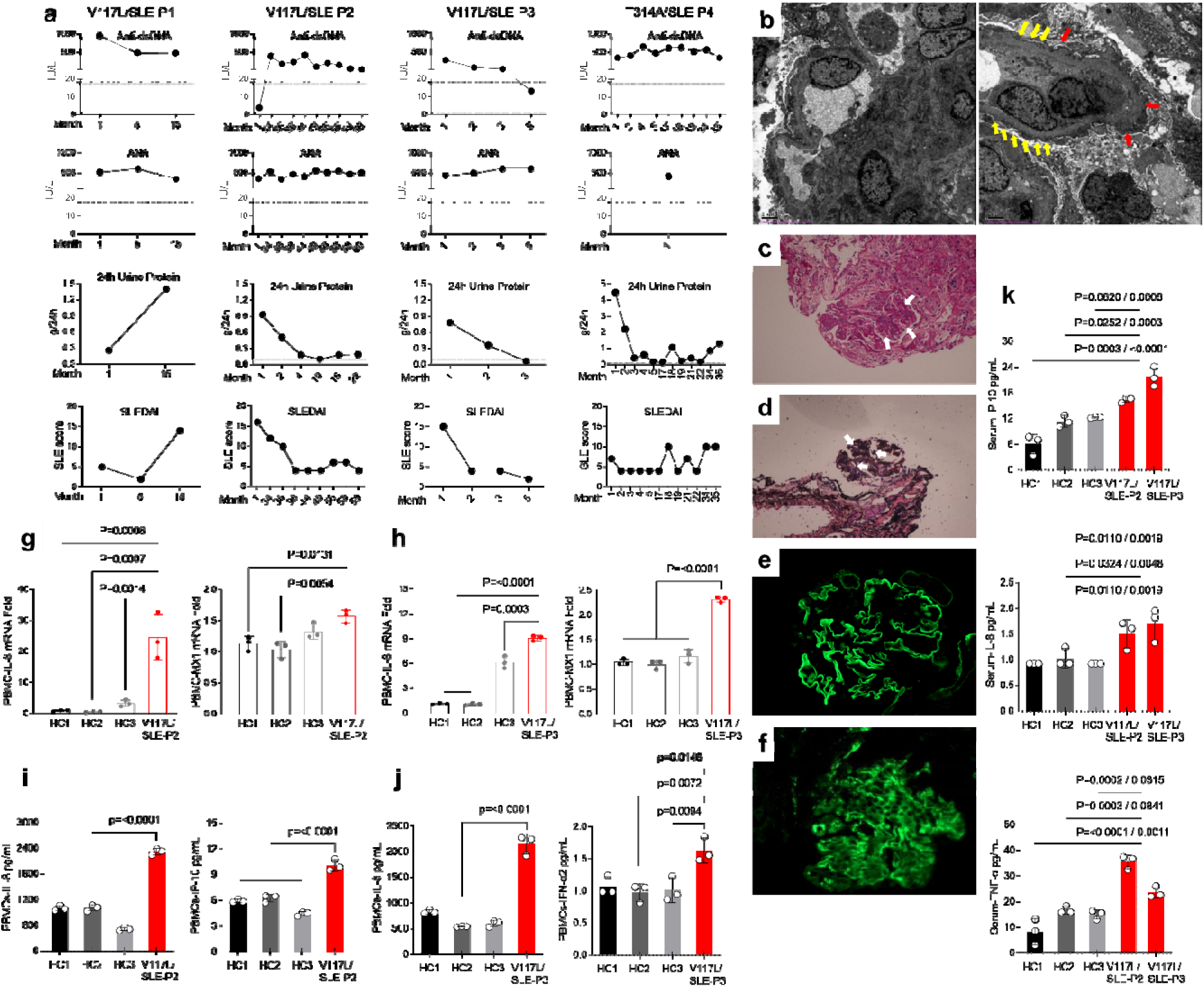
Clinical characteristics related to UNC93B1 V117L and T314A variants. (a) Anti-dsDNA autoantibodies, ANA autoantibodies, 24hrs. urine protein, and SLEDAI score of disease for P1, P2, P3 (all V117L) and P4 (T314A). Data is for monthly visits post diagnosis. (b-f) Kidney pathology for patient with UNC93B1 T314A. (b) Transmissio Electron Microscopy showed 2 glomeruli, capillary endothelial cell proliferation, obvious vacuolar degeneration, re blood cells, monocytes, and neutrophil aggregation in little vascular loops. The podocyte foot process has diffuse effacement (yellow arrows), with the cell swelled and cavitated. The basement membrane has diffuse thickening, with thickness up to 1300 nm (red arrows). Mesangial cells are aggregated. Electron densification was deposited in stromal hyperplasia, the subepithelial, intrabasement, subendothelial and mesangial zones. In tubulo-interstitium, epithelial cells were cavitated with a small amount of renal tubular atrophy and inflammatory cells infiltrated into the renal interstitium. In renal interstitial vessels, there was red blood cell aggregation in the loops of little capillaries an thickening of the arteriole wall, P4. (c) Periodic Acid–Schiff (PAS) stain shows thickening of the basement membrane, and mesangial and endothelial cell proliferation (white arrows), P4. (d) PAS methenamine (PASM) stain showed thickening of the basement membrane, and a small number of platinum ear like structures (white arrows), P4. (e) Immunofluorescence for IV collagen of the glomerular basement membrane showed α3:+ve, α5+ve, P4. (f) Immunofluorescence of kidney tissue showing complement 3 deposition, P4. (g) Quantitative RT-PCR analysis of IL-8 and MX1 mRNA expression in the PBMCs of P2 compared to healthy controls. (h) Quantitative RT-PCR analysis of IL-8 and MX1 mRNA expression in the PBMCs of P3 compared to healthy controls. (i) Production of IL-8 and IP-10 in the supernatant of PBMCs isolated from P2 and healthy controls measured by CBA. (j) Production of IL-8 and IFNα2 in the supernatant of PBMCs isolated from P3 and healthy controls measured by CBA. (k) Levels of IP-10, IL-8, and TNFα in the plasma from P2 and P3 compared to healthy controls. Indicated p values were determined by two-way ANOVA.

## UNC93B1 variants drive inflammation and immune dysfunction via TLR7-IRAK1/4

To study the direct effect of these rare UNC93B1 variants we created an in vitro model with THP-1 monocytes overexpressing WT, V117L or T314A. The patient mutations trigger spontaneous upregulation of transcripts for IFNβ and the interferon stimulated genes, IFIT3, ISG-15, and ISG20L2 (**Fig. 3a**). Upregulated mRNA levels of inflammatory cytokines, IL-8, IL12a, and TNFα were also observed (**Fig. 3a**). In addition, the mutated UNC93B1 induced phosphorylation of IRF5, NFκB, and MAPK (JNK1,2,3 and P38) as shown by western blotting (**Fig. 3b**). Overexpression of UNC93B1 V117L, and T314A resulted in excess secretion of IFNα, IFNβ, and IL-6 as detected by ELISA (**Fig. 3c**) and IL-8 and IP-10 as detected by cytokine bead array (**Fig. 3d**). For an unbiased comparison, we performed RNA sequencing in cells overexpressing UNC93B1 V117L or T314A. From the most significantly upregulated genes, at least six are known biomarkers for disease activity in lupus (DEFB1 ^15^, PRLR ^16^, S100A8/9 ^17^, FCER2 ^18^, PRG2 ^19^) (**Supp Fig. 3a and 3d**). Further gene set and pathway analysis pointed towards programs related to innate immune response, phagosome activity, and antigen processing and presentation (**Fig. 3e,f and Supp Fig. 3b,c,e,f**). Collectively, these results indicate that the UNC93B1 variants identified can intrinsically promote inflammation and immune dysfunction associated with type I IFN and NF-kB signaling pathways.

**Fig. 3.**
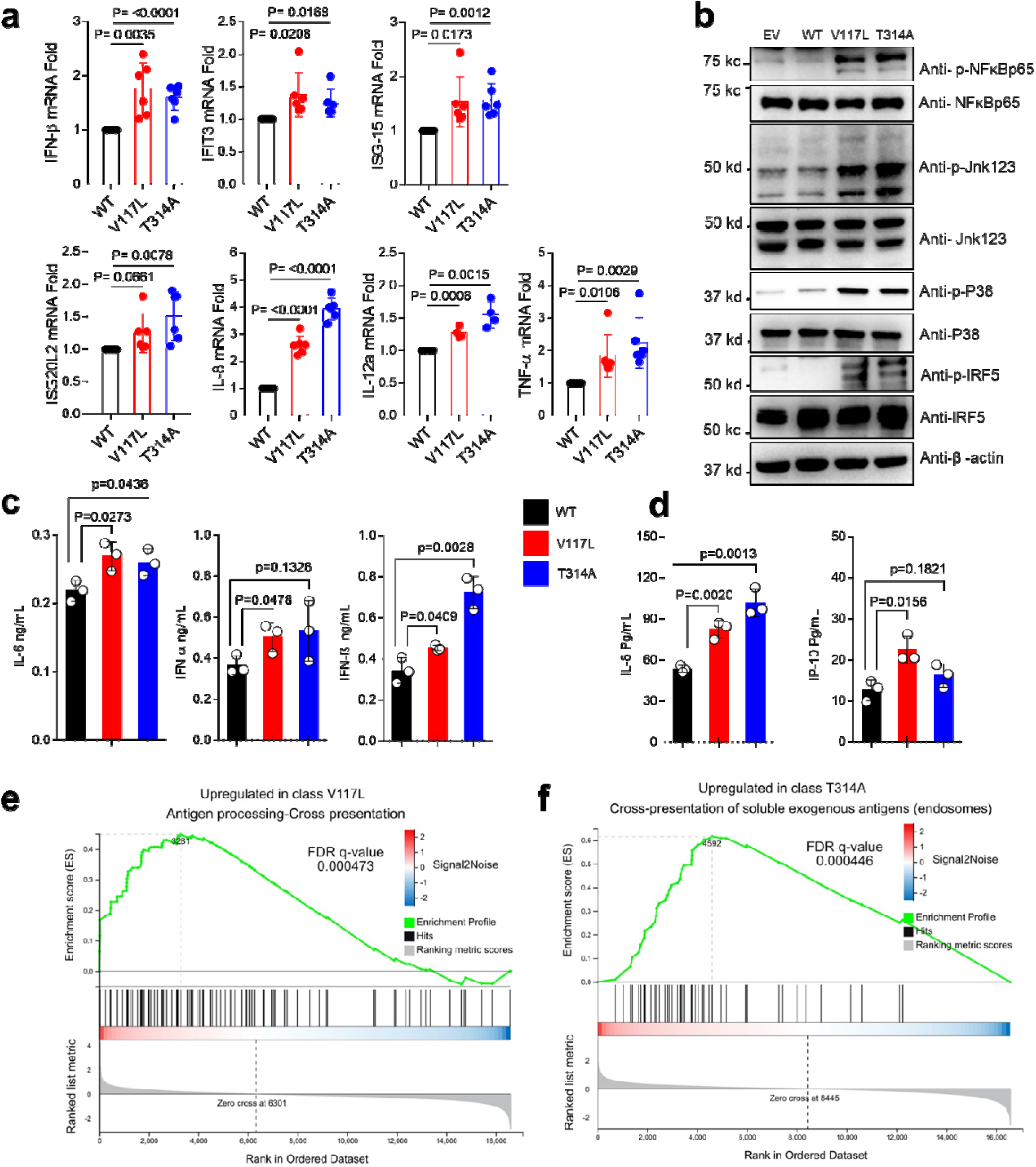
V117L and T314A UNC93B1 variants spontaneously induce IFN and NF-kB signaling pathways in THP-1 cells. (a) Quantitative RT-PCR analysis of IFNβ, IFIT3, IL-8, IL-12A, ISG-15, ISG20L2, and TNFα expression in the indicated THP-1 cell lines, n=3 biological replicates. Three independent experiments. (b) Levels of phosphorylated NFκB, JNK1,2,3, p38, and IRF5, as measured by immunoblot, in lysates of the indicated THP-1 cells. Data are representative of three independent experiments. (c) Production of IL-6, IFNα, and IFNβ in the indicated THP-1 cell lines. n=3, three independent experiments (ELISA). (d) Production of IP-10 and IL-8 in the indicated THP-1 cell lines. n=3 biological replicates (CBA). Indicated p values were determined by unpaired t-test. (e,f) Gene Set Enrichment Analysis (GESA) with WT as control groups and UNC93B1^V117L^ or UNC93B1^T314A^ as treatment groups which shows a significant association for genes relating to antigen processing and cross presentation. mRNA was extracted from THP-1 of UNC93B1^WT^, UNC93B1^V117L^, and UNC93B1^T314A^, n=3 biological replicates.

Some of the most attractive therapeutic targets downstream of UNC93B1 and the nucleic acid sensing TLRs (NAS-TLRs) are the signaling adaptor kinases IRAK1 and IRAK4 ^20^. A dual targeting inhibitor was tested to confirm that this is the pathway triggered by UNC93B1 V117L and T314A, and assess potential clinical utility. Transcriptionally, IRAK1/4 inhibition was efficacious, returning IL-8, IL-12a, TNF-α, and the interferon-related genes, IFN-β, ISG20L2, and ISG-15 close to baseline (**Fig. 4a**). The pathway was confirmed by western blot of cell lysates, where induced phosphorylation of IRF5, NFκB, and MAPK (JNK1,2,3 and P38) was also downregulated due to IRAK1/4 inhibition (**Fig. 4b**). In agreement, the downstream cytokines produced by UNC93B1 V117L and T314A Thp-1 cells (eg. IL-8, TNF-α, IFN-β, and IP-10) were significantly blunted (**Fig 4c, d**). These findings point towards IRAK1/4 as the dominant signaling pathway triggered by variants in UNC93B1 found in patients with childhood-onset SLE. As IRAK1/4 are adaptors for NAS-TLR signaling, we next investigated which could be involved in this phenotype by stimulating patient PBMCs using Poly I:C, R848, and CPG-C, the ligands of TLR3, TLR7/8, and TLR9 respectively. The results revealed that patient PBMCs stimulated by R848 showed induced genes expression of IFIT1 and TNF-α more than healthy controls, but not when stimulated by Poly I:C and CPG-C (**Fig 4e**). In addition, secretion of TNF-α, IL-1β, and IL-6 in the supernatant of PBMCs of P2 were increased specifically after stimulation by R848, but not Poly I:C or CPG-C (**Fig 4f**). These results were independently confirmed using PBMCs of P3 (**Fig 4g**). Taken together, genetic variation in UNC93B1 regulates inflammation in patients with childhood-onset SLE through a TLR7 – IRAK1/4 pathway.

**Fig. 4.**
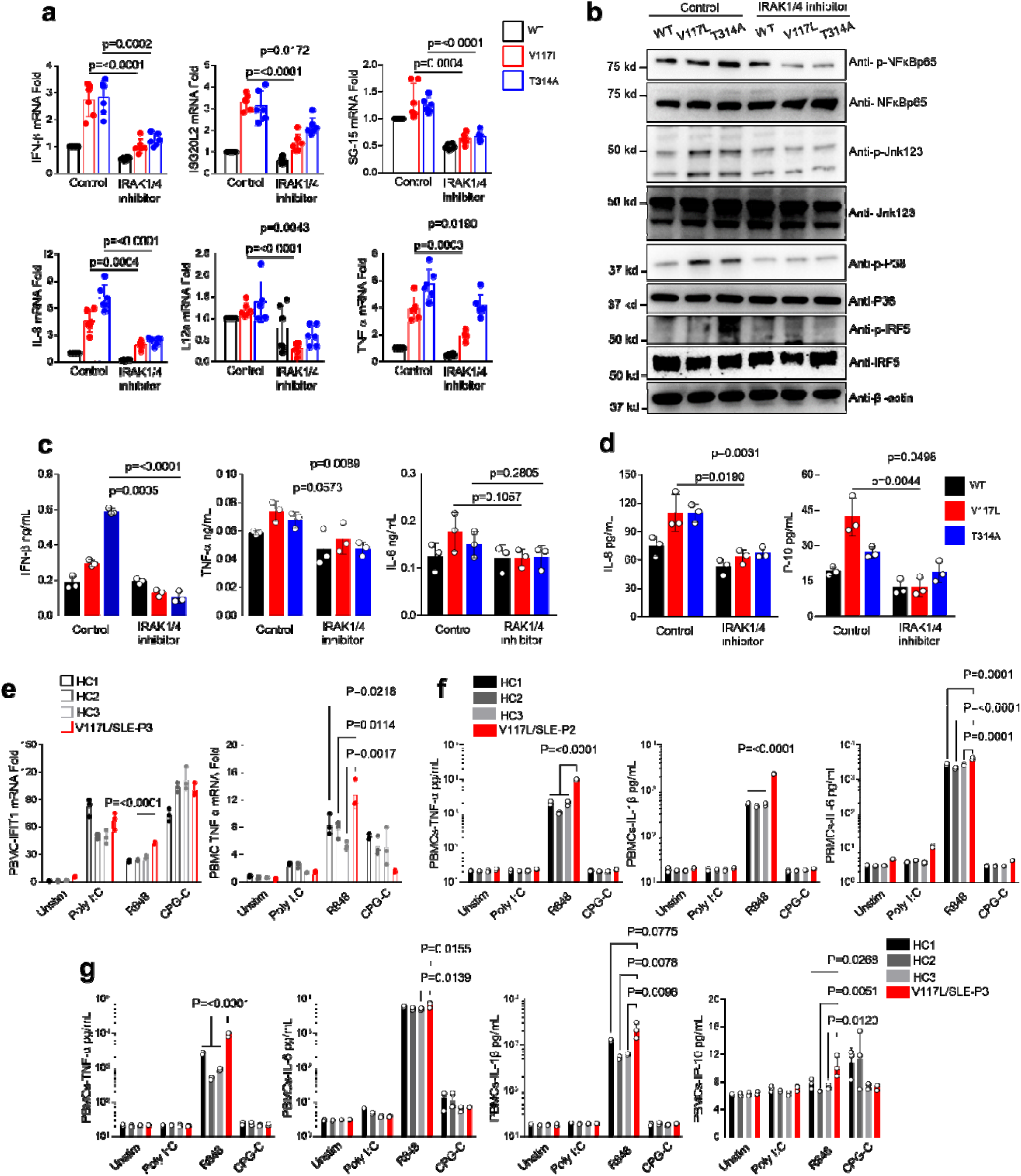
UNC93B1 genetic variation drive inflammation via TLR7/IRAK1/4 axis. (a) Quantitative RT-PCR analysis of IFNβ, ISG20L2, ISG-15, IL-8, IL-12a, and TNFα expression in the indicated THP-1 cell lines after incubation with IRAK1/4 inhibitor for 24hrs, n=3 biological replicates. Data are representative of two independent experiments. (b) Levels of phosphorylated NFκB, JNK1,2,3, p38, and irf5 as measured by immunoblot using lysates of the indicated THP-1 cells after incubation with IRAK1/4 inhibitor for 24hrs. (c) Production of IFNβ and TNFα in the indicated THP-1 cell lines after incubation with IRAK1/4 inhibitor for 72hrs. n=3 biological replicates. (d) Production of IL-8 and IP-10 in the indicated THP-1 cell lines after incubation with IRAK1/4 inhibitor for 72hrs. n=3 biological replicates (CBA). Indicated p values in a, c, and d were determined by unpaired t-test. (e) Quantitative RT-PCR analysis of IFIT1 and TNF-α mRNA expression in the PBMCs of P3 compared to healthy controls after stimulation by 10μg HMW Poly I:C, 1μg R848, and 4μg CPG-C for 12hrs. Indicated p values were determined by two-way ANOVA. (f) Production of TNF-α, IL-1β, and IL-6 in the supernatant of PBMCs of P2 compared to healthy controls after stimulation by 10μg/mL HMW Poly I:C, 1μg/mL R848, and 4μg/mL CPG-C for 8hrs. Indicated p values were determined by two-way ANOVA. (g) Production of TNF-α, IL-6, IL-1β and IP-10 in the supernatant of PBMCs of P3 compared to healthy controls after stimulation by 10μg/mL HMW Poly I:C, 1μg/mL R848, and 4μg/mL CPG-C for 12hrs. Indicated p values were determined by two-way ANOVA.

## Lupus-like disease in mice with a mutation orthologous to UNC93B1 V117L

Given that UNC93B1 (V117L) is a highly significant risk factor for childhood-onset SLE, but also present in the general population, we sought to confirm pathogenicity in vivo and created mice with the orthologous mutation V138L. Heterozygous and homozygous UNC93B1^V138L^ mice were born at normal Mendelian ratios and initially appear healthy, however they lose weight (**Fig. 5a**), develop splenomegaly with increased spleen cellularity (**Fig. 5b**) and have a low kidney size (**Fig. 5c**). In addition, lupus-associated serum anti-dsDNA and anti-smith autoantibodies were increased in knock-in mice (**Fig. 5d**), but no change was noted for total immunoglobulin G (data not shown). Bone marrow populations of CD45^+^ immune cells, macrophages, and CD11b^+^/Gr-1^+^ cells were increased along with CD16^+^/CD14^-^ cells, regulatory T cells, and activated T cells (**Fig. 5e**). Functionally, the intracellular production of IFN-γ in the immune cells of bone marrow was also upregulated in the mutant mice (**Fig. 5f**). In the spleen of UNC93B1^V138L^ mice there were increased B cells and a decreased CD4 T cell /B cell ratio, along with elevated regulatory T cells, activated T cells, germinal center B cells and CD45^+^/CD4^+^/CD44^+^ cells (**Fig. 5g**). Serum cytokine analysis demonstrated that UNC93B1^V138L^ leads to upregulation of IL-12p70, IP-10, IL-6 and IFN-γ (**Fig. 6a**). Histologically, there was overt pathology in kidney, spleen, lung, and pancreas where disease scores of mesangial expansion, extramedullary hematopoietic cell hyperplasia and inflammatory cell infiltrate, respectively, were elevated in mice with the UNC93B1^V138L^ variant (**Fig. 6b**). Molecularly, increased mRNA of the interferon-related genes, MX1, ISG20L2, IFIT1, and IRF-7 were observed in kidney tissue homogenate (**Fig. 6c**).

**Fig. 5.**
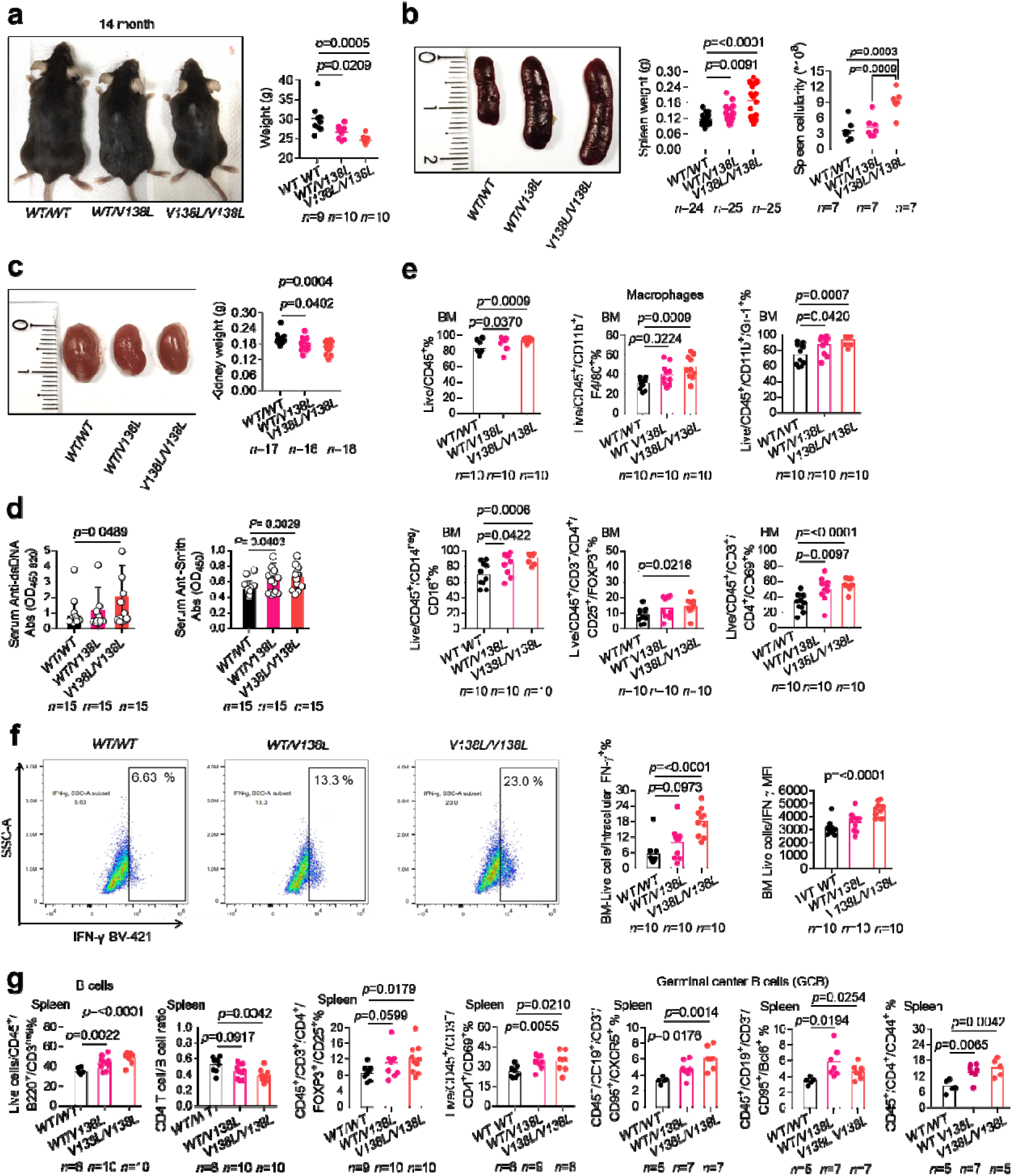
UNC93B1^V138L^ mice develop lupus-like disease. (a) Appearance and weight of UNC93B1^WT/WT^, UNC93B1^WT/V138L^, and UNC93B1^V138L/V138L^ mice. (b) Spleen weight and splenocyte count of indicated mice. Mice were age matched 8-14 months old, the data pooled from three independent experiments (c) Kidney weight of indicated mice. Mice were age matched 8-14 months old, the data pooled from two independent experiments. (d) Serum autoantibodies to dsDNA or Smith for indicated mice (n=15). Mice were age matched 8-14 months old, the data pooled from three independent experiments. (e) Flow cytometric analysis of indicated immune cell populations in bone marrow (BM) of UNC93B1^WT/WT^, UNC93B1^WT/V138L^, and UNC93B1^V138L/V138L^ mice (n=indicated). (f) Intracellular cytokine staining of IFN-γ in BM immune cells of indicated mice (n=10). (g) Flow cytometric analysis of indicated immune cell populations in spleen of UNC93B1^WT/WT^, UNC93B1^WT/V138L^, and UNC93B1^V138L/V138L^ mice (n=indicated). Mice were age matched 8 months (for B cells, CD4 T cell /B cell ratio, regulatory T cells, and activated T cells) or 14 months old (for germinal center B cells and CD45^+^/CD4^+^/CD44^+^ cells).

**Fig. 6.**
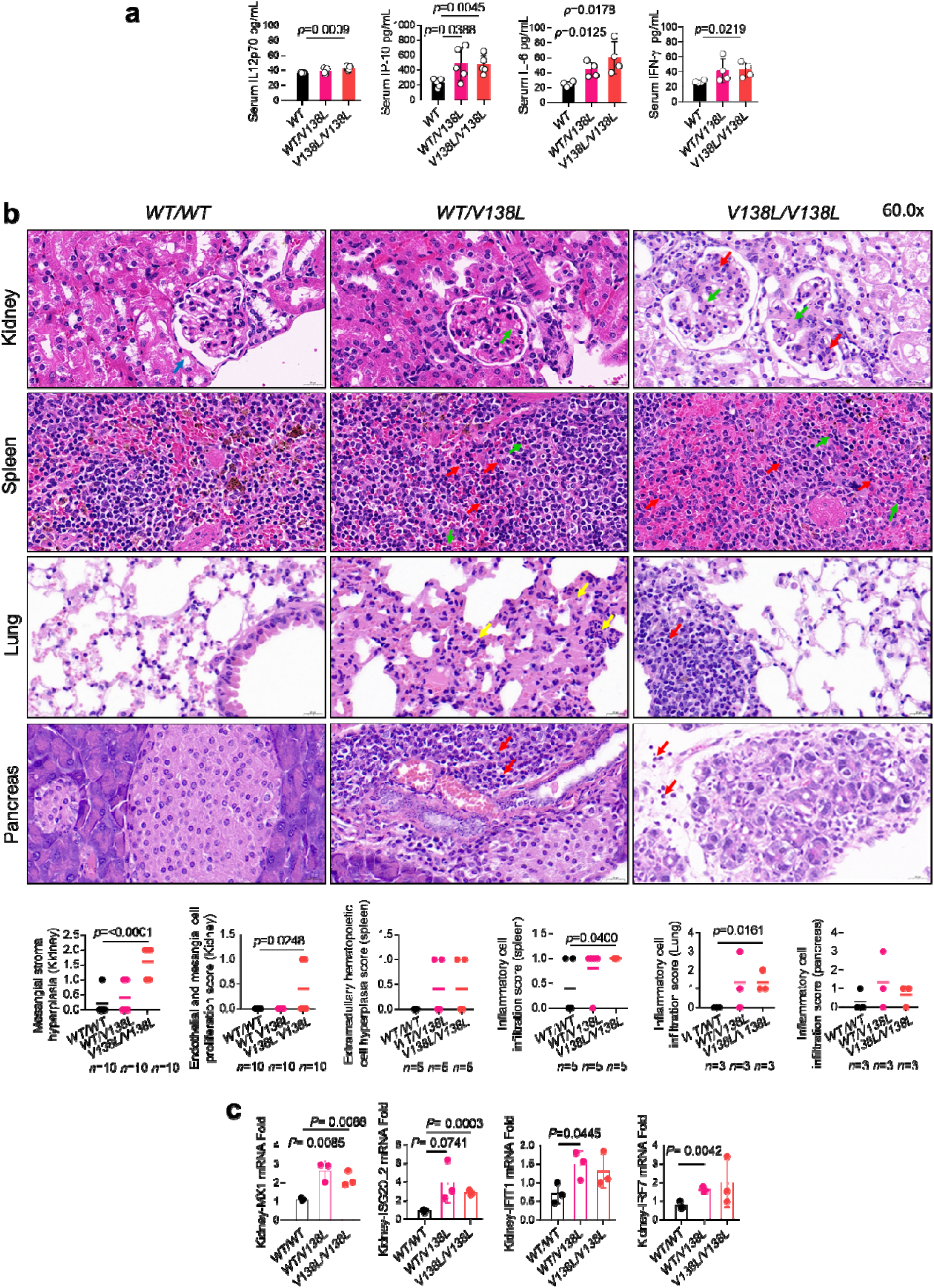
UNC93B1^V138L^ mice develop systemic inflammation and organ damage. (a) Serum IL-12p70, IP-10, IL-6, and IFN-γ of indicated mice (n=5 for IL-12p70 and IP-10; n=4 for IL-6, and IFN-γ). For IP-10 mice were 8 months old and for IL-12p70, IL-6, and IFN-γ 10 months old. (b) Haematoxylin and eosin (H&E) staining of the kidneys from indicated mice (n=10), the green arrows indicated mesangial stroma hyperplasia, and the red arrows indicated endothelial and mesangial proliferation. Mice were age matched 8-14 months, the data pooled from three independent experiments. H&E staining of the spleens from indicated mice (n=5), the green arrows indicated the extramedullary hematopoietic cells and the red arrows indicated the inflammatory cells. Mice were age matched 8 months. H&E staining of the lungs from indicated mice (n=3), the yellow arrows (granulocytes) and red arrow (lymphocytes) indicated inflammatory cells infiltration, mice were 14 months old. H&E staining of the pancreases from indicated mice (n=3), the red arrows indicate inflammatory cells infiltration (granulocytes or lymphocytes), mice were 14 months old. The graphs show the pathological disease scores. (c) Quantitative RT-PCR analysis of MX1, ISG20L2, IFIT1 and IRF7 mRNA in kidney tissues from indicated mice (n=3). Mice were age matched 8 months old. Statistical analysis was done using unpaired *t*-tests. The exact *p* values are shown.

Bone marrow-derived macrophages (BMDM) isolated from UNC93B1^V138L^ mice experienced increased mRNA of the interferon-related genes, IRF-7 and IFIT1 when compared to BMDM of UNC93B1^WT^ mice (**Fig. 7a**). Meanwhile, phosphorylation of the UNC93B1/TLR7 signaling pathway, including IRF5, NFκB, and MAPK (JNK1,2,3 and P38) was also observed to be activated in UNC93B1^V138L^ BMDM (**Fig. 7b**). Additionaly, UNC93B1^V138L^ BMDM exhibit elevated intrinsic intracellular production of TNF-α (**Supp Fig. 4a**). Unbiased RNA sequencing indicates that all of the most highly upregulated genes are known to be inducible by type I or type II IFN ^21^ and includes the central TLR signaling molecule IRF7, as well as IRF3 and TBK1 ^22^ (**Supp Fig. 4b, c**). Consistently, this was associated with pathway analysis implicating TLR signaling, TNF signaling, antigen processing and presentation (**Supp Fig. 3d-f**). Overall, there is a very significant association with genes listed in KEGG disease: Systemic Lupus Erythematosus (**Fig. 7c**). To investigate the specificity of NAS-TLR signalling in this mouse model, BMDMs were stimulated by TLR ligands and tested for inflammatory markers. As in human samples, data from these experiments showed the involvement of TLR7 in mouse lupus-like phenotype. IFIT1, IRF7, ISG-15, and TNF-α genes were more highly expressed in BMDM of mice with the UNC93B1^V138L^ variant after stimulation with R848 but not poly I:C, or CPG-C (**Fig. 7d**). Also, secretion of CXCL1, IP-10, and CCL5 in the supernatant of UNC93B1^V138L^ BMDM were elevated after R848 stimulation but not after stimulation by poly I:C, or CPG-C (**Fig. 7e**). These findings implicate enhanced TLR7 signalling as the pathway driving inflammation and a lupus-like phenotype in mice with the equivalent gene variant to humans contributing to childhood-onset SLE.

**Fig. 7.**
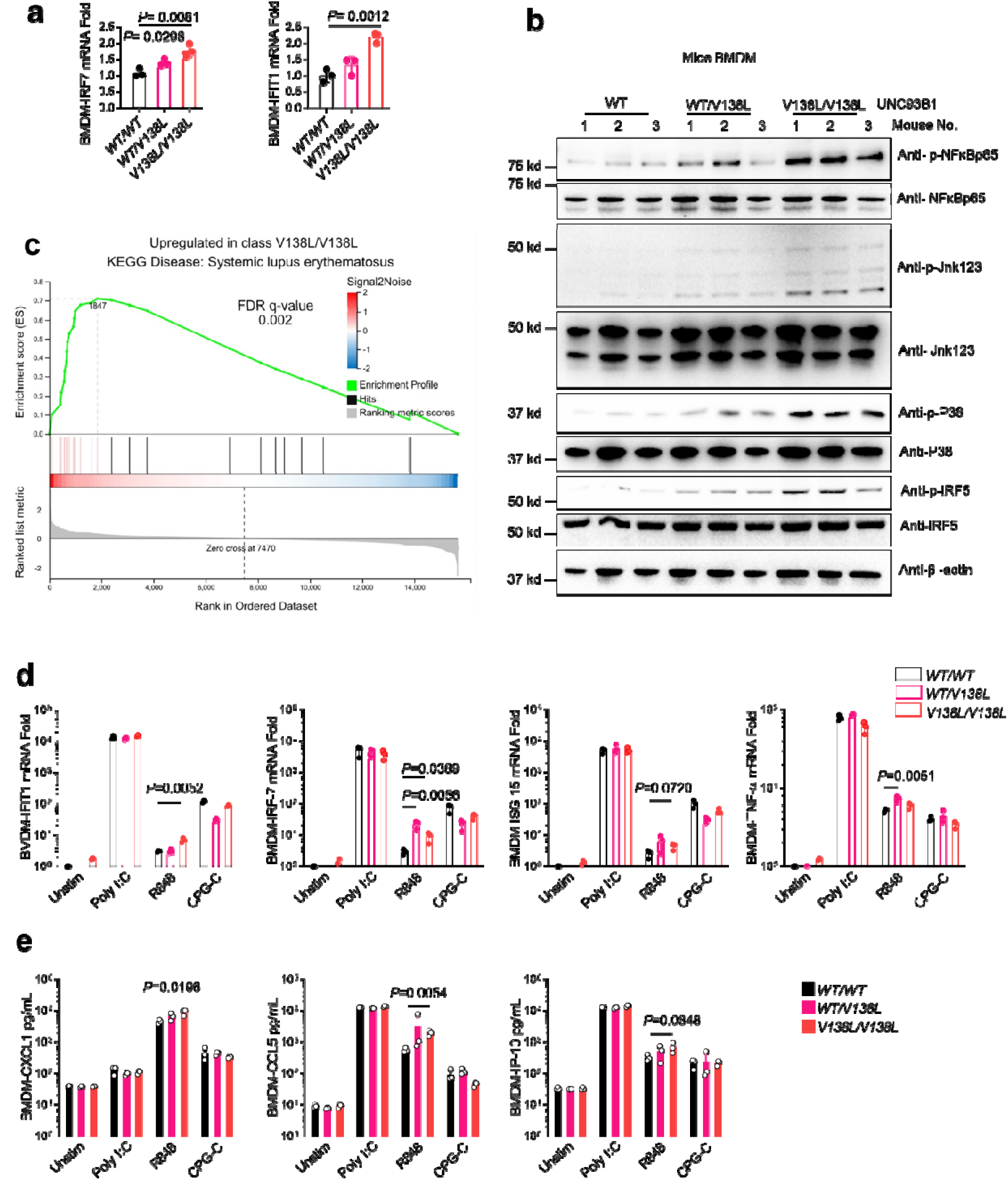
UNC93B1^V138L^ drives increased inflammation and TLR7 responses in mice. (a) Quantitative RT-PCR analysis of IRF7 and IFIT1 mRNA in bone marrow-derived macrophage (BMDM)from indicated mice (n=3). Mice were age matched 8 months. Statistical analysis was done using unpaired *t*-tests. The exact *p* values are shown. (b) Levels of phosphorylated NF-κB, JNK, P38, and IRF5 as measured by immunoblot, in lysates of BMDM of indicated mice (n=3). Mice were age matched 8 months. (c) RNA sequencing was performed for UNC93B1^V138L^ BMDM compared to control, n=3 biological replicates. Mice were age matched 8 months. Gene Set Enrichment Analysis significantly associated with Systemic Lupus Erythematosus. (d) Quantitative RT-PCR analysis of IFIT1, IRF-7, ISG-15 and TNFα mRNA expression in the BMDM from indicated mice (n=3) after stimulation by 40μg/mL HMW Poly I:C, 2μg/mL R848, and 10μg/mL CPG-C for 24hrs. Mice were age matched 14 months old. Statistical analysis was done using unpaired t-tests. The exact p values are shown. (e) Production of CXCL1, CCL5, and IP-10 in the supernatant of mice BMDM isolated from indicated mice (n=3). Measured by CBA after stimulation by 40μg/mL HMW Poly I:C, 2μg/mL R848, and 10μg/mL CPG-C for 24hrs. Mice were age matched 14 months old. Statistical analysis was done using unpaired t-tests. The exact p values are shown.

Therefore, UNC93B1^V138L^ drives a lupus-like disease in mice at a cellular level, with relevant end organ damage. This is associated with activation of the TLR7 signaling pathway and inflammatory genes expression that is consistent with the clinical presentation of patients with the orthologous genetic change, who suffer from childhood-onset SLE.

## Discussion

Our discovery of rare genetic changes in UNC93B1 that predispose to childhood-onset SLE was facilitated by a large body of work culminating in the discovery of lupus-causing variants in TLR7^4^. Interestingly, all patients with UNC93B1 variants identified in this study were female, which could relate to a lack of TLR7 X-chromosome inactivation ^23^ however could also reflect the strong gender predisposition of this disease in general. We found that the spontaneous inflammatory signaling via IRAK1/4, due to the variants in UNC93B1 identified here, is associated with increased responses to stimulation of TLR7/8. This seems logical based on the current literature, however further work is required for formally delete TLR7 and resolve pathology from the UNC93B1^V138L^ mouse model we generated. Theoretically, there should also be an endogenous ligand to stimulate TLR7 in this context, which could be guanosine or other nucleic acid ^24^. Furthermore, although the structural location of UNC93B1 T314A presents a logical mechanism to impact TLR7, the molecular effect of UNC93B1 V117L is not yet clear. Mechanistic insight into this process will be extremely useful, and potentially clinically actionable in the future as TLR7 inhibitors are being developed ^25^.

Not only are TLR7 inhibitors in clinical trials, but also IRAK1/4 inhibitors could have therapeutic benefit for the patients identified, based on our results. So far, none of the patients characterised in this study have gone on to receive JAK inhibitors or biological therapeutics that would also target type I IFN signaling. Those approaches should be beneficial, and this could be particularly important given that UNC93B1 V117L is present in the general East Asian population, where it could be a significant cause of SLE ^26^. Within this population there is a strong geographical bias (**Fig. 1c**) for which the underlying basis is unknown. Given that the family members of the affected individuals in this study were all apparently healthy, we currently consider that UNC93B1 V117L is not a monogenic disease-causing allele with incomplete penetrance, but rather a very strong risk factor to develop childhood-onset SLE.

Our mouse model represents an important confirmation of the patient findings, and shows that a gene dosage effect of the gain-of-function variant is present. Although no homozygous humans have been identified so far, we can speculate that they may have more severe, or greater likelihood to develop, disease. As UNC93B1 V117L is present in the general population as a rare but highly significant risk factor for disease, the preclinical efficacy of new therapeutic modalities can be accurately modelled for the resulting patient population using our mice avatars. The mouse model should also be useful to determine the cellular contribution of UNC93B1 to lupus-like disease, either intrinsically in B-cells, or with activation of innate immune signaling from antigen presenting cells, and the distinct contribution of type I IFNs compared to other inflammatory programs. We would expect this to be similar to other gain-of-function UNC93B1 mouse models of lupus that are published ^27^.

Overall, these findings are not just the first patients with childhood-onset SLE due to variants in UNC93B1, but they bridge the gap from rare monogenic diseases to show that this signaling pathway is relevant for the incidence of disease more generally. Moreover, therapeutics targeting this specific pathway are being developed and can be tested first in a mouse model for which there is a corresponding patient population.

## Data Availability

RNAseq datasets are submitted and the identifier will be included in the manuscript when published. All other data is available in the main text or the supplementary materials.

## Acknowledgments

We are grateful to all members of the Zhang and Masters laboratories for supplying reagents and technical advice or assistance. We also thank Salah Adlat for his kind help during drafting this manuscript.

## Author contributions

S.L.M. and Yuxia.Z. conceived the project; M.A. conducted all experiments with assistance from I.E.; M.A. performed data analysis, drafted figures and wrote the first version of manuscript; K.H-S. performed structural analysis; J.Z. assisted with animal experiments; M.L., H.X., M.Y, B.L., Z.Z., Y.L., J.C. were involved in experiments and data analysis; Z. L. performed bioinformatics analysis; J.C., X.L., C.G., assisted with provision of human samples; P.Z., X.G., were involved in clinical management; Yan.Z., was involved in WES sequencing analysis; S.L.M. supervised the project; M.A. wrote the manuscript with input from all authors.

## List of Supplementary Materials

Materials and Methods

Figures S1-S5

Table S1-S3

