## Supplementary Data for "Genetic variants in UNC93B1 predispose to childhood-onset systemic lupus erythematosus"

### Methods and Materials

#### Patients

Female patients (P) 1, 2, 3, and 4 at ages between 5-10 presented symptoms of systemic inflammation associated with lupus nephritis. Patients blood and other samples were collected at Guangzhou Women and Children's Medical Centre, Guangzhou, China. All patients were diagnosed as SLE patients according to EULAR/ACR classification criteria (**Fig. 2a-f, Supp Fig. 1a-c, and Supp Table 2**). Written informed consent has been obtained from the authorized individual for all participated patients. The Guangzhou Women and Children's Medical Centre Medical Ethics Committee approved the study procedures ([2021]073B00) and ([2021]240A01) in consistence with Helsinki Declaration about ethics in using human samples.

#### DNA sequencing

Whole Exome sequencing and sanger sequencing was performed at Novogene. The frequency of V117L in the East Asian population was obtained from PGC.Han 2.0 (*1*).

#### hPBMCs isolation and plasma separation

Non-coagulated blood samples from the SLE patients and healthy controls were diluted in equal volume of phosphate buffered saline (PBS), and dispensed slowly and gently along the side of a 15ml conical tube containing 2ml of Ficoll-Hypaque density gradient. This was followed by centrifugation at 800g for 20 minutes, 4 ACC, 4 DEC, 4 C$^o$. The cloud-like cell layer of cells was collected from the interphase into a new tube, and washed twice with sterile PBS by centrifuging at 1000 RPM for 5 minutes. The pellet was resuspended in complete culture medium consisting 90% RPMI-1640, 10% fetal bovine serum (FBS), and 100 U/ml/100µg/ml P/S and then incubated at 37°C, 5% $CO_2$ and 95% humidity. The Human peripheral blood mononuclear cells (hPBMCs) were used in experiments immediately after isolation. For plasma separation, non-coagulated blood samples from the SLE patients and healthy controls were centrifuged at 1500g/10 minutes then the upper yellow fluid was separated into new tube by pipetting without disturbing the layer of buffy coat. Samples stored at -40C$^o$ up to the time of analysis.

#### Antibodies and reagents

For immunoblots the following antibodies were used: anti-FLAG (F1804, Sigma-Aldrich), anti-NF-κB (82425, Cell Signaling), anti-Phospho-NF-κB (Ser468) (3039S, Cell Signaling), anti-jnk123 (ab179461, Abcam), anti- Phospho-jnk123 (ab124956, Abcam), anti-P38 (#8690 , Cell Signaling), anti-Phospho-P38 (9211S, Cell Signaling), anti-IRF5 (#20261, Cell Signaling), anti-Phospho- IRF5 (Ser437) (#PA5-64760, Invitrogen), anti-β-actin (AC026, ABclonal), anti-rabbit IgG, HRP-linked antibody (7074), and anti-mouse IgG, HRP-linked antibody (7076). For flow cytometry the following antibodies and reagents were used: anti-TNF-α (506324, Biolegend), anti-IFN-γ (505830, Biolegend), anti-CD45 (103132, Biolegend), Zombie Aqua-A (Biolegend), anti-F4/80 (123137, Biolegend), anti-CD11b (101205, Biolegend), anti-CD14 (123335, Biolegend), anti-CD16 (158004, Biolegend), anti-CD3 (100248, Biolegend), anti-CD4 (100406, Biolegend), anti-FOXP3 (126407, Biolegend), anti-CD25 (102043, Biolegend), anti-CD44 (103059, Biolegend), anti-CD69 (104507 or 104510, Biolegend), anti-Gr-1 (108416, Biolegend), anti-CD19 (159812, Biolegend), anti-CD95 (152612, Biolegend), anti-Bcl-6 (358510, Biolegend), anti-CD45R/B220 (103222 or 103284, Biolegend), CXCR5 (145517, Biolegend), and fixation and permeabilization kit (88-8824-00, Thermofisher). For TLRs stimulation, the following ligands were used: HMW Poly I:C (tlrl-pic, InvivoGen), and R848 (tlrl-r848, InvivoGen) used in human and mouse experiments, CPG-C (tlrl-m362, InvivoGen) used in mouse experiments, and CPG-C, ODN 2395 VacciGrade (vac-2395-1, InvivoGen), used in mice BMDM stimulation.

**Mice**

Animal studies were approved by Institutional Animal Care and Use Committee of Guangzhou Medical University (GY2022-035). Mice lines (*UNC93B1*$^{WT/V138L}$ and *UNC93B1*$^{V138L/V138L}$) were designed and developed by Shanghai Model Organisms Center, Inc (Shanghai, China). Generation of the point mutation mice model at exon3 (*$^{138}$V to L*) of *UNC93B1* gene via CRISPR/Cas9 technology. Briefly, Cas9 mRNA and gRNA were produced by in vitro transcription; oligo donor DNA was synthesized; the mixture of Cas9 mRNA, gRNA and donor DNA was microinjected into fertilized eggs (C57BL/6J), then three positive F0 mice were identified by PCR and sequencing; F0 mice were crossed with wild type C57BL/6J mice to generate F1 mice, then four positive F1 mice were identified by PCR and sequencing. The guide RNA used was: GTGTAGAGCAGGGCAGCGAT. The Knock-in locus: GCCTCCTGCAGATGCAACTGATCCTGCACTATGATGAGACCTACAGAGAGGTGAAG TATGGCAACATGGGGCTGCCGGACATCGATAGCAAGATGCTGATGGGTATCAACGT

GACGCCTATCGCTGCCCTGCTCTACACACCTGTGCTCATCAG (The underlined letter is GuideRNA target site and the blue font is mutant site).

The oligo donor DNA sequence is: GTATGGCAACATGGGGCTGCCGGACATCGATAGCAAGATGCTGATGGGTATCAAC**C**TGACGCCTATAGCTGCCCTGCTCTACACACCTGTGCTCATCAGGTGCCAAACTTCTGTTTCCGCGCC (The proposed mutation is highlighted in bold red, silent mutation is highlighted in green). All mice used for analysis in this project are females.

**UNC93B1 lentivirus constructs and transduction**

For human UNC93B1 (NM_030930) overexpression, constructs of wild type and mutated, V117L and T314A UNC93B1 were inserted into Ubi-MCS-3FLAG-SV40-Cherry-IRES-puromycin. Site-directed mutagenesis technology was used to generate c.G349T:p.V117L and c.A940G:p.T314A mutation using wild type UNC93B1 construct. These plasmids along with PSPAX2 and pMD2G were used to produce lentiviruses in supernatant. Designing and synthesis of plasmids, and lentiviruses production were done at Genechem Laboratory, Shanghai, China. THP-1 cells were transduced by indicated lentiviruses to generate stable cell lines overexpressing UNC93B1 WT, V117L, and T314A. 2 µg/mL puromycin (Santa Cruz Biotechnology) treatment and FACS sorting by BD FACSAria lll were performed for positive cell selection lasting two weeks after infection. The infection efficiency and stable overexpression were verified by western blotting assay for flag expression and flow cytometry for mCherry by CYTEK NL-CLC.

**Cell culture**

THP-1 cells were obtained from American Type Culture Collection (ATCC) and cultured accordingly. THP-1 and human PBMCs were cultured in RPMI 1640 medium supplemented with 10% FBS, 2mM L-glutamine, and 1% Penicillin-Streptomycin. All cells were cultured at 37°C with 5% $CO_2$ and 100% humidity. THP-1 incubated with 5µM IRAK1/4 inhibitor I (15409, Sigma-Aldrich) as indicated in results section.

**mRNA extraction and qPCR**

Extraction of mRNA from THP-1, BMDM, or kidney tissue homogenate was performed using EZ-press RNA Purification Kit and converted to cDNA using the 4× Reverse Transcription Master

Mix Kit (with gDNA Remover) from EZ Bioscience and 1000ng mRNA/sample according to the manufacturer's instructions. The quantification of RNA concentration was done using VARIOSKAN LUX (Thermo SCIENTIFIC). Quantitative real-time PCR was conducted using the 2× Color SYBR Green qPCR Master Mix (ROX2 plus) from EZ Bioscience, cDNA, RNase Free $dH_2O$, and primers shown in Supplementary Table 3 by LightCycler 480 ll (96 or 384), Roche according to the manufacturer's instructions.

**Western blotting**

Cell lysates were extracted using a low-salt lysis buffer (50mM HEPES pH 7.5, 150mM NaCl, 1mM EDTA, 1.5mM MgCl2, 10% glycerol, 1% Triton X-100) supplemented with 5mg/ml protease and phosphatase inhibitors cocktail. Protein quantification performed using Pierce BCA Protein Assay Kit (23227, Thermofisher) according to manufacturer's instructions. 10 or 12 % sodium dodecyl sulfate-polyacrylamide gel electrophoresis (SDS-PAGE) was used for protein lysates (20 μg) with loading buffer and then transferred to PVDF (Millipore Co., USA). Membranes were blocked with 5% skimmed milk in tris-buffered saline tween-20, TBST, and then incubated overnight at 4°C with primary antibodies. After washing, the membranes were incubated for 1h at room temperature with secondary antibody conjugated with HRP. Finally, TBST-washed membranes were treated with enhanced chemiluminescent (ECL) for detection (Bio-Rad, USA). Developed membranes were imaged using the Image Lab detection system (Bio-Rad, USA).

**ELISA and CBA**

Enzyme-linked immunosorbent assay (ELISA) and cytokines bead array (CBA) experiments were carried out using serum/plasma or/and cell supernatants according to the manufacturer's instructions. ELISA kits for human, IFNα (CSB-E08636h), IFNβ (CSB-E09889h), IL-6 (CSB-E04638h) and TNFα (CSB-E04740h) were purchased from Cusabio. ELISA kits for mouse, LBIS mouse anti-dsDNA (637-02691, FUJIFILM Wako Shibayagi Corporation), mouse anti-Sm Ig's (total (A+G+M) (5405, ALPHA DIAGNOSTIC INTERNATIONAL), and mouse IgG (6320, ALPHA DIAGNOSTIC INTERNATIONAL) were purchased as indicated. Absorbance were measured using VARIOSKAN LUX (Thermo SCIENTIFIC). Human (740390) and Mouse (740621) LEGENDplex™ 13-plex CBA anti-Virus Response Panel kits from Biolegend were used in cytokines quantification using V-bottom 96-plate protocol. Beads were analyzed by CYTEK NL-CLC. Data were collected using CYTEK NL-CLC software, SpectroFlo and analyzed using the LEGENDplex™ Data Analysis Software Suite (BioLegend).

**Separation of mice serum**

Mice orbital whole blood samples were collected into tubes without anticoagulant and incubated at room temperature for at least 15 minutes to get blood coagulated. The coagulated blood samples were centrifuged at 1500g/10 minutes then the upper yellow fluid was separated into new tube by pipetting. Samples stored at -40C$^{o}$ up to the time of analysis.

**Tissue digestion and flow cytometry**

The whole spleen was minced into a tiny pieces in 6-well plate on ice and filtered by 70 µm strainer using plunger end of the syringe with rinsing by 5mL PBS with 2% FBS. The collected tissue filtrate were centrifuged at 500g /5 minutes at 4 C$^{o}$. The cells pallet suspended in RBC lysis buffer and incubated for 5 minutes at room temperature and then mixed with double volume of PBS with 2% FBS. The immune cells were collected in the pellet after centrifugation at 500g /5 minutes at 4 C$^{o}$.

Collected bone marrow from mouse femur bones by centrifuging using technique of two layers of tubes were incubated with RBC lysis buffer for 5 minutes at room temperature and then mixed with double volume of PBS with 2% FBS. The immune cells were collected in the pellet after centrifugation at 500g /5 minutes at 4 C$^{o}$.

For flow cytometry, 1 million cells were incubated with Zombie Aqua-A (Biolegend) for 15 min in dark, then washed to be stained by antibodies for cell surface markers and incubated for 30 min at 4C$^{o}$ and then washed. Fixation and permeabilization kit (88-8824-00, Thermofisher) was used for fixation and intracellular permeablization for intracellular and nuclear markers according to manufacturer's protocol. Cells analyzed by CYTEK NL-CLC. Data were collected using CYTEK NL-CLC software, SpectroFlo and analyzed using FlwoJo_v10.8.1. Gating strategy shown in (**Supp Fig. 5**).

**RNA sequencing**

THP-1 cells or mice BMDMs were suspended in trizol and sent to Beijing Genomic Institute (BGI) laboratory for RNA sequencing. Analysis was performed using in-house BGI software or using published methodology (*2-4*).

**Isolation of mouse bone marrow-derived macrophage**

The bone marrow of mice femur bones was collected aseptically. BMDMs were generated by incubating the bone marrow immune cells in a complete medium, RPMI 1640, 10% FBS , 2mM L-glutamine, and 1% Penicillin-Streptomycin containing 50ng/mL macrophage colony stimulating factor (M-CSF) (Peprotech) for 6-8 days at 37°C with 5% $CO_2$ and 100% humidity.

**Intracellular staining for TNFα and IFNγ**

BMDMs or bone marrow immune cells were stimulated by 1X cocktail of phorbol 12-myristate 13-acetate (PMA), ionomycin, brefeldin A and monensin from Invitrogen™ ( 00-4975-93) for 4-6 hrs at 37°C with 5% CO2 and 100% humidity and then stained by PE-Cy7 TNFα and BV-421 IFNγ antibody using fixation and permeabilization kit (88-8824-00, Thermofisher) according to manufacturer's protocol. Cells analyzed by CYTEK NL-CLC. Data were collected using CYTEK NL-CLC software, SpectroFlo and analyzed using FlwoJo_v10.8.1.

**Histopathology**

The kidney, spleen, lung, and pancreas tissues were preserved in 4% paraformaldehyde. Tissue processing and histopathology reporting was performed by Wuhan Servicebio Technology Laboratory. For disease scoring (blinded) of tissues sections stained by H&E, 0=within the normal range, 1= Very slight (The changes that appear just exceeded the normal range), 2= slight (Lesions may be observed but not severe), 3=medium (Lesions are obvious and are likely to be more severe), 4= severe (lesions have taken up the entire tissue and organs). For kidney pathology of mesangial stroma and proliferation, the score calculated for around 50 glomeruli per mouse.

**Statistical Analysis**

Statistical analysis was performed using Prism 7 software (GraphPad Software Inc.). Unpaired t-test and One-way or Two-way ANOVA were used to compare differences between and groups. A p-value less than 0.05 was considered significant.


**Supplementary Table 1**

Incidence of UNC93B1 (V117L) in South Coast Han with and without childhood-onset SLE

***p*** Probability value = $< 0.05$ (significant)

**OR 95 % CI:** 6.54 to 53.5

| Genotype | V117L | WT | Total | Chi-square | p-value | Odds Ratio |
|---|---|---|---|---|---|---|
| Childhood-onset SLE | 7 | 265 | 272 | 50.18 | <0.0001 | 17.9 |
| South Coast Han | 6 | 4057 | 4063 | | | |
| Total | 13 | 4322 | 4335 | | | |

#### Supplementary Table 2

Clinical parameters for patients with UNC93B1 variants

| Clinical presentation | P1 (V117L) | P2 (V117L) | P3 (V117L) | P4 (T314A) |
|---|---|---|---|---|
| Fatigue | - | - | + | - |
| Fever | + | - | + | + |
| Alopecia | - | + | + | - |
| Mucocutaneous involvement | + | + | + | + |
| Cardiac involvement | + | - | + | - |
| Vascular manifestations | + | - | - | - |
| Renal involvement | + | + | + | + |
| Gastrointestinal involvement | + | + | + | - |
| Pulmonary involvement | + | - | + | - |
| Hematologic abnormalities | + | - | + | + |
| Ophthalmologic involvement | - | - | - | + |
| **Laboratory tests** | | | | |
| Anti-Sm | + | - | - | - |
| Anti-Ro/SSA | - | + | + | + |
| Anti-La/SSB | - | - | - | - |
| Anti-nRNP | + | + | - | - |
| Hematuria | +++ | ++++ | +++ | +++ |
| Proteinuria | ++ | ++++ | ++++ | ++ |
| Urine WBC | ++ | + | + | - |
| **Therapy (chronological)** | | | | |
| | MSS | OS | BED | MSS |
| | OS | MSS | TM | OS |
| | CCP | OH | VD | PS |
| | GG | CG | CG | MM |
| | LT | HS | PS | VD |
| | PS | VD | MM | CG |
| | VD | GG | | CF |
| | MTX | THY | | GSG |
| | FA | CPP | | FS |
| | | MM | | DD |
| | | BM | | CPP |
| | | DS | | GRL |
| | | BFG | | LT |
| | | GOG | | |

MSS, methylprednisolone sodium succinate; OS, omeprazole sodium; CPP, cyclophosphamide; GG, glutamine granules; LT, loratadine; PS, prednisone; MTX, methotrexate; FA, folic acid; OH, ondansetron hydrochloride; CG, compound glycyrrhizin; HS, hydroxychloroquine sulfate; THY, thymosin; BM, Belimumab; DS, dexamethasone sodium; BFG, bovine basic fibroblast growth factor eye gel; GOG, Ganciclovir Ophthalmic Gel; BED, brinzolamide eye drops; TM, timolol maleate eye drops; CF, compound falcodine; MM, mycophenolate mofetil; VD, Vitamin D; GSG, L-glutamine sodium gualenate; FS, fosinopril sodium; DD, Dipyridamole; GRL, Glucuronolactone.

**Supplementary Table 3**

Primers used for QPCR

| Gene | Species | Forward Primer | Reverse Primer |
|---|---|---|---|
| IFNB1 | Human | AGTAGGCGACACTGTTCGTG | AGCCTCCCATTCAATTGCCA |
| IFIT3 | Human | AAAAGCCCAACAACCCAGAAT | CGTATTGGTTATCAGGACTCAGC |
| ISG15 | Human | CGCAGATCACCCAGAAGATCG | TTCGTCGCATTTGTCCACCA |
| ISG20L2 | Human | AGGAAATGCCAAGCACCGAAA | TGAAGGTTCAGAGTGCAACTTAG |
| IL-8 | Human | TTTTGCCAAGGAGTGCTAAAGA | AACCCTCTGCACCCAGTTTTC |
| IL12A | Human | CCTTGCACTTCTGAAGAGATTGA | ACAGGGCCATCATAAAAGAGGT |
| TNF-α | Human | CCTCTCTCTAATCAGCCCTCTG | GAGGACCTGGGAGTAGATGAG |
| MX1 | Human | GTTTCCGAAGTGGACATCGCA | CTGCACAGGTTGTTCTCAGC |
| IFIT1 | Human | TTGATGACGATGAAATGCCTGA | CAGGTCACCAGACTCCTCAC |
| GAPDH | Human | GGAGCGAGATCCCTCCAAAAT | GGCTGTTGTCATACTTCTCATGG |
| B2M | Human | AGCAGCATCATGGAGGTTTG | AGCCCTCCTAGAGCTACCTG |
| IRF-7 | Mouse | AAGCTGGAGCCATGGGTATG | CGATGTCTTCGTAGAGACTGTTGG |
| IFIT1 | Mouse | AGAGTCAAGGCAGGTTTCTG | TGTGAAGTGACATCTCAGCTG |
| MX1 | Mouse | GATCCGACTTCACTTCCAGATGG | CATCTCAGTGGTAGTCAACCC |
| ISG20L2 | Mouse | ACCAACTTGGAGGCCTATGG | AGGGGTCAGCCAAGACAATG |
| ISG15 | Mouse | CGATTTCCTGGTGTCCGTGA | AGCCAGAACTGGTCTTCGTG |
| TNF-α | Mouse | CCAAATGGCCTCCCTCTCAT | TGGTGGTTTGCTACGACGTG |
| GAPDH | Mouse | ATCAAGAAGGTGGTGAAGCA | AGACAACCTGGTCCTCAGTGT |

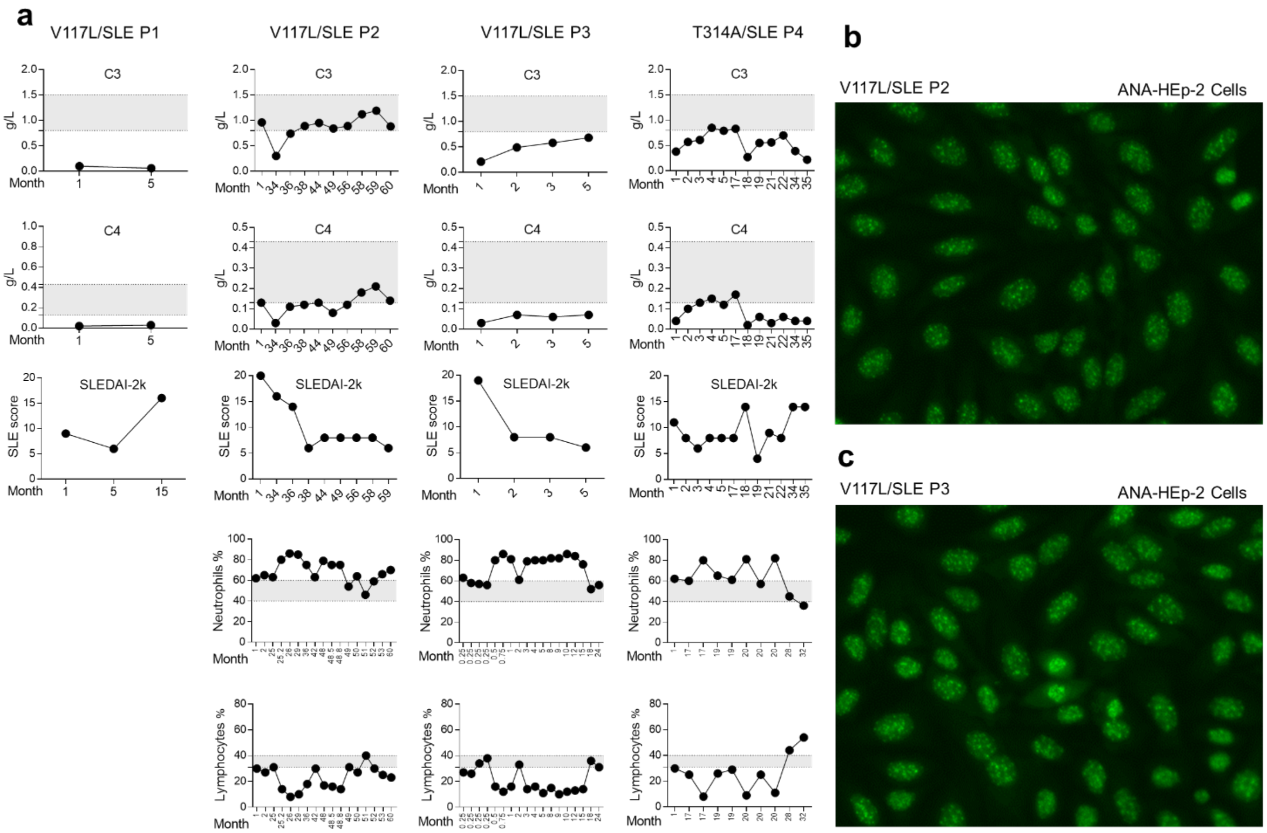


**Supplementary Fig. 1. Clinical characteristics associated with UNC93B1 V117L and T314A.** (a) Complement 3/4 levels, SLE score (SLEDAI-2K), circulating neutrophils and lymphocytes counts for patients with UNC93B1 V117L or T314A. Data is for monthly visits post diagnosis. (b, c) ANA autoantibodies by HEp-2 cells for P2 and P3 showing mixed patterns in which the speckled pattern is predominant.

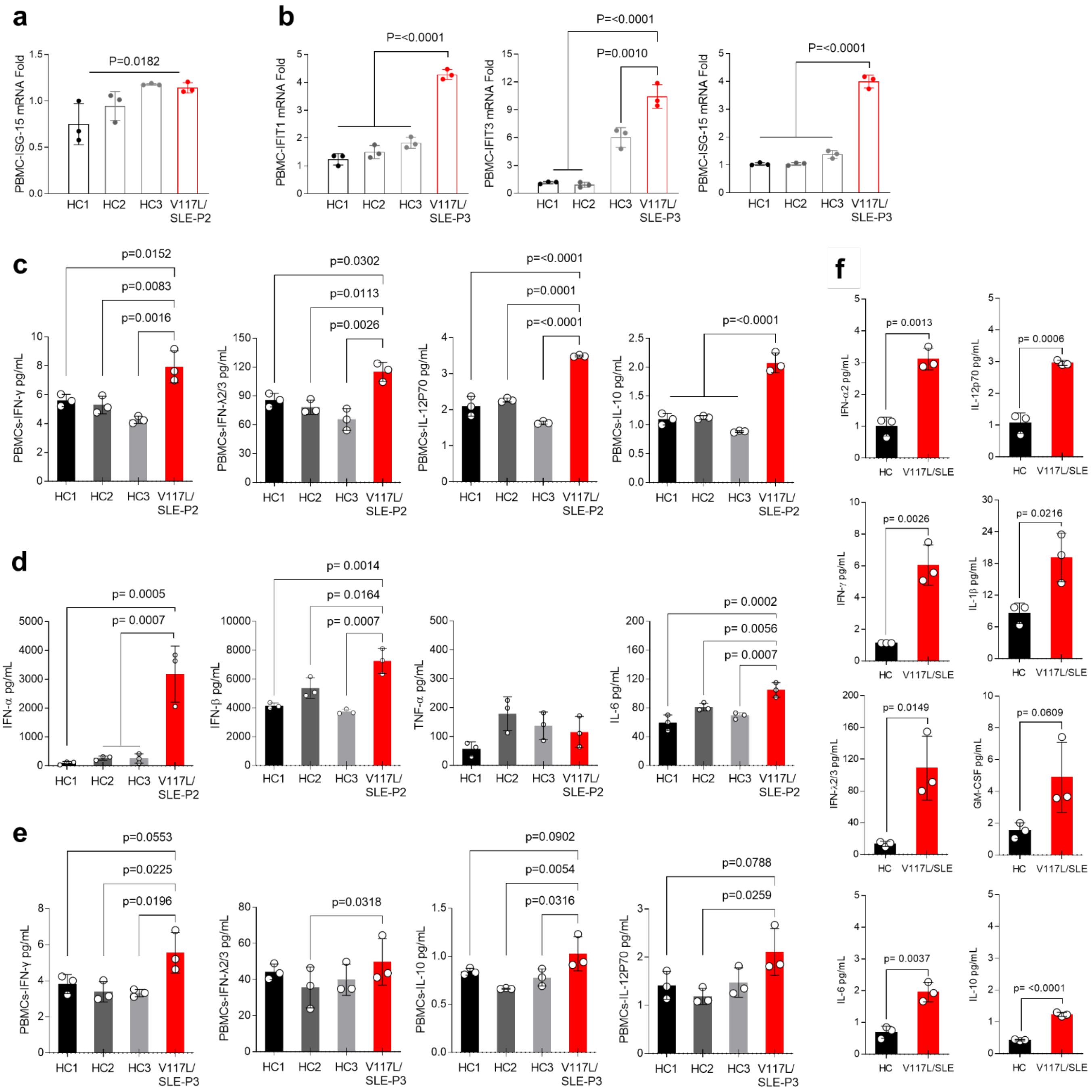


**Supplementary Fig. 2. Patients with UNC93B1 V117L display elevated interferons and inflammatory cytokines.** (a) Quantitative RT-PCR analysis of ISG-15 mRNA expression in the PBMCs of P2 compared to healthy controls. p value was determined by one-way ANOVA. (b) Quantitative RT-PCR analysis of IFIT1, IFIT3, and ISG-15 mRNA expression in the PBMCs of P3 compared to healthy controls. (c) Production of IFNγ, IFNλ2/3, IL-12P70, and IL-10 in the supernatant of PBMCs isolated from P2 and healthy controls measured by CBA. (d) Production of IFNα, IFNβ, TNFα, and IL-6 in the supernatant of PBMCs isolated from P2 and healthy controls measured by ELISA. (e) Production of IFNγ, IFNλ2/3, IL-10, and IL-12P70 in the supernatant of PBMCs isolated from P3 and healthy controls measured by CBA. Indicated p values in b–e were determined by two-way ANOVA. (f) Levels of IFN- α2, IFNγ,

IFN$\lambda$2/3, IL-6, IL-12P70, IL-1$\beta$, GM-CSF, and IL-10 in the plasma from P2 compared to healthy controls measured by CBA, p values were determined by unpaired t-test.

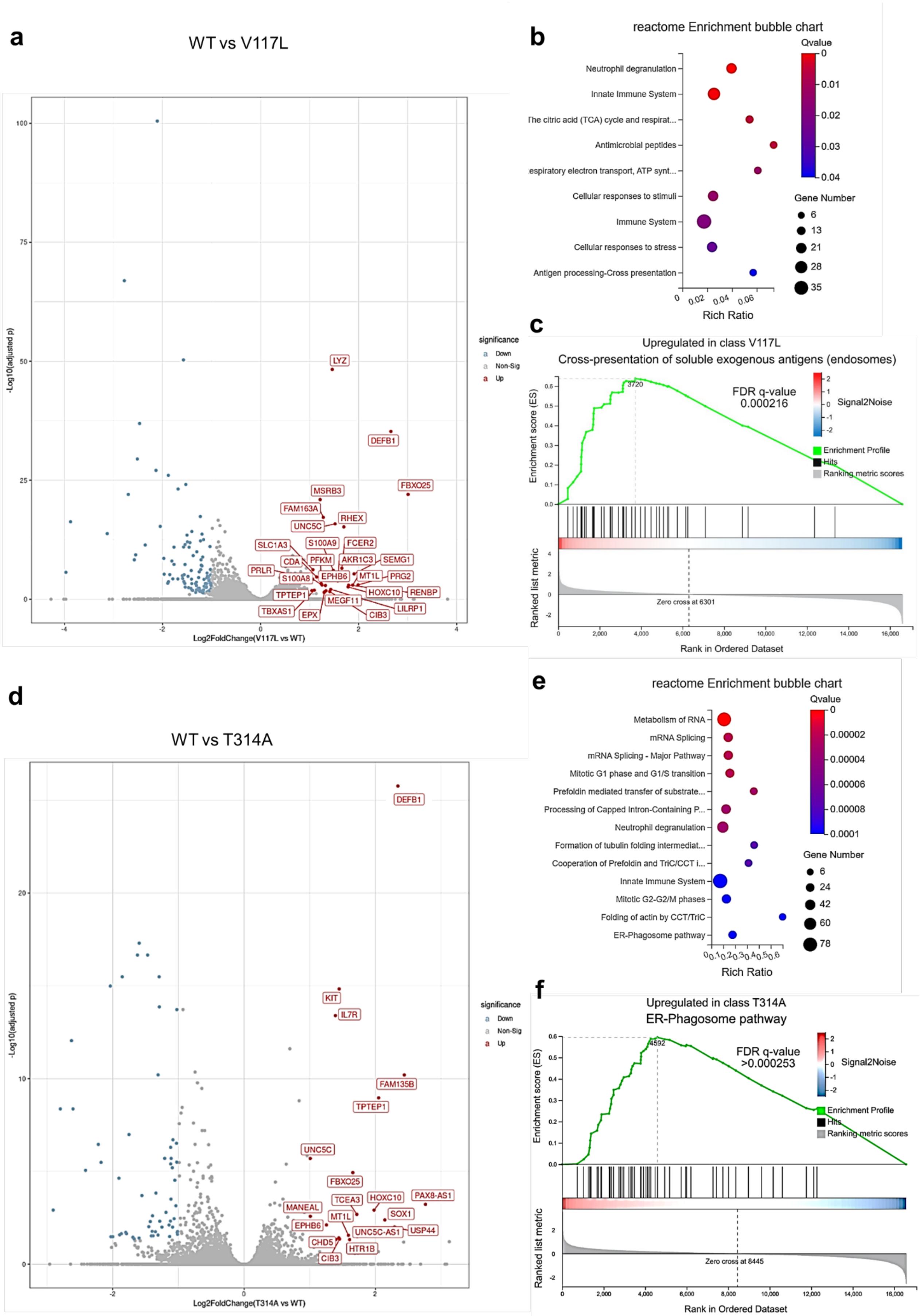
a
WT vs V117L
-Log10(adjusted p)
Log2FoldChange(V117L vs WT)
significance
Down
Non-Sig
Up
LYZ
DEFB1
FBXO25
MSRB3
FAM163A
RHEX
UNC5C
SLC1A3
S100A9
FCER2
CDA
PFKM
AKR1C3
SEMG1
PRLR
S100A8
EPHB6
MT1L
PRG2
HOXC10
RENBP
TPTEP1
MEGF11
LILRP1
TBXAS1
EPX
CIB3
b
reactome Enrichment bubble chart
Neutrophil degranulation
Innate Immune System
The citric acid (TCA) cycle and respirat...
Antimicrobial peptides
espiratory electron transport, ATP synt...
Cellular responses to stimuli
Immune System
Cellular responses to stress
Antigen processing-Cross presentation
Rich Ratio
Qvalue
Gene Number
c
Upregulated in class V117L
Cross-presentation of soluble exogenous antigens (endosomes)
FDR q-value
0.000216
Signal2Noise
Enrichment Profile
Hits
Ranking metric scores
Enrichment score (ES)
Ranked list metric
Zero cross at 6301
Rank in Ordered Dataset
d
WT vs T314A
-Log10(adjusted p)
Log2FoldChange(T314A vs WT)
significance
Down
Non-Sig
Up
DEFB1
KIT
IL7R
FAM135B
TPTEP1
UNC5C
FBXO25
TCEA3
HOXC10
PAX8-AS1
MANEAL
MT1L
SOX1
EPHB6
UNC5C-AS1
USP44
CHD5
HTR1B
CIB3
e
reactome Enrichment bubble chart
Metabolism of RNA
mRNA Splicing
mRNA Splicing - Major Pathway
Mitotic G1 phase and G1/S transition
Prefoldin mediated transfer of substrate...
Processing of Capped Intron-Containing P...
Neutrophil degranulation
Formation of tubulin folding intermediat...
Cooperation of Prefoldin and TriC/CCT i...
Innate Immune System
Mitotic G2-G2/M phases
Folding of actin by CCT/TriC
ER-Phagosome pathway
Rich Ratio
Qvalue
Gene Number
f
Upregulated in class T314A
ER-Phagosome pathway
FDR q-value
>0.000253
Signal2Noise
Enrichment Profile
Hits
Ranking metric scores
Enrichment score (ES)
Ranked list metric
Zero cross at 8445
Rank in Ordered Dataset

**Supplementary Fig. 3. DEGs and pathway analysis of inflammation in UNC93B1$^{V117L}$ and UNC93B1$^{T314A}$ THP-1 cells.** (a-c) RNA sequencing was performed for UNC93B1$^{V117L}$ THP-1 cells compared to control, n=3 biological replicates. (a) Differentially expressed genes (DEGs) presented as a volcano plot. (b) Reactome enrichment analysis of significantly upregulated genes highlights innate immune system and antigen processing/presentation. (c) Gene Set Enrichment Analysis also significantly associated with antigen processing/presentation. (d-f) RNA sequencing was performed for UNC93B1$^{T314A}$ THP-1 cells compared to control, n=3 biological replicates. (d) DEGs presented as a volcano plot. (e) Reactome enrichment analysis of significantly upregulated genes highlights innate immune system and ER/Phagosome pathways. (f) Gene Set Enrichment Analysis also significantly associated with ER/Phagosome function.

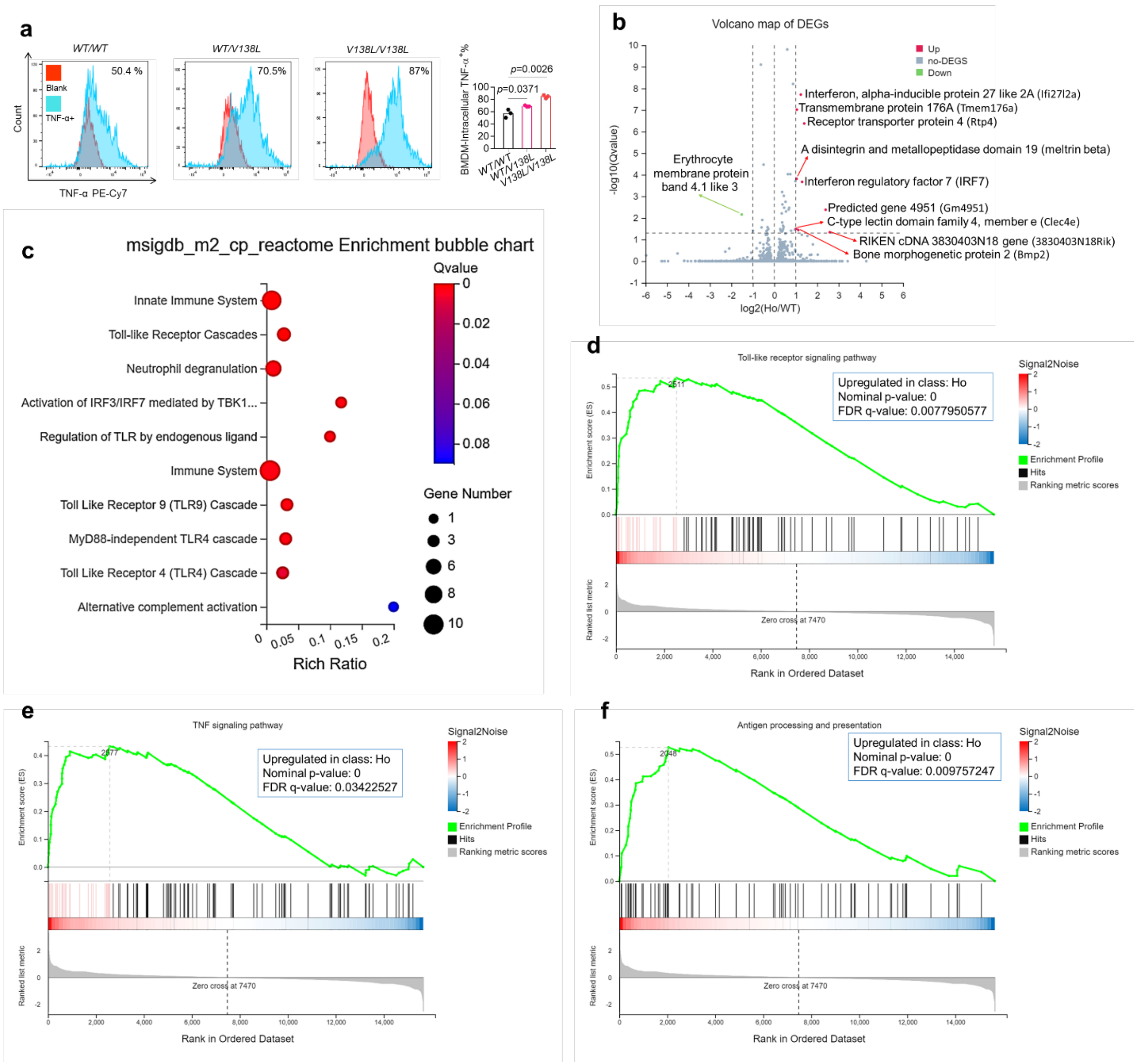


**Supplementary Fig. 4. DEGs, and upregulated pathways of inflammation in UNC93B1$^{V138L}$ homozygous mice compared to wild type.** (a) Intracellular cytokine staining of TNF-α in bone marrow-derived macrophage (BMDM) of indicated mice (n=3). Red histograms are unstained controls. (b-f) RNA sequencing was performed for UNC93B1$^{V138L}$ BMDM compared to control, n=3 biological replicates. (b) Differentially expressed genes presented as a volcano plot. (c) msigdb_m2_reactome enrichment analysis of significantly upregulated genes highlights innate immune system and toll-like receptor pathways. (d-f) Gene Set Enrichment Analysis significantly associated with TLR signaling, TNF signaling, and antigen processing/presentation. Mice were 8 months old.

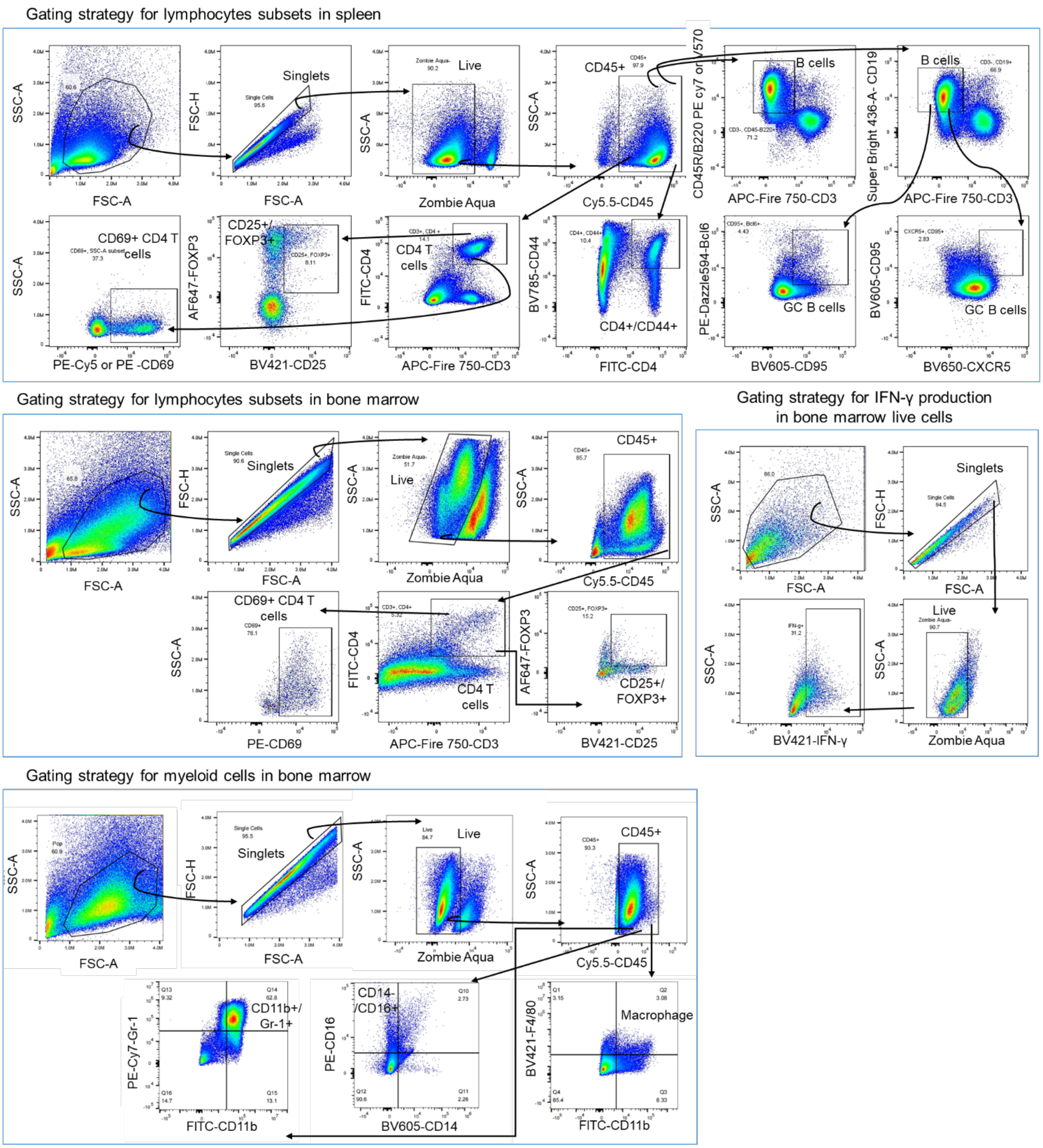


**Supplementary Fig. 5. Gating strategies.** Representative gating strategies for lymphocytes subsets, B cells, CD4 T cells, T regulatory cells, activated T cells, or/and germinal center B cells in bone marrow or/and spleen, myeloid cells in bone marrow, macrophage, monocytes, inflammatory Gr-1$^{+}$ cells, and IFN-γ producing cells. These strategies used for data presented in figure 5.
